# Validation of the AndroWash Automated System for Standardized and Efficient Sperm Preparation in Assisted Reproductive Technology

**DOI:** 10.64898/2025.11.29.25341236

**Authors:** Sohan Sahu, Vikram Rajput, H. M. Mishra, Bhupendr Sahu

## Abstract

Sperm preparation is a critical, yet highly operator-dependent, step in Assisted Reproductive Technology (ART) workflows, often leading to variability in Total Motile Sperm Count (TMSC) recovery and prolonged processing times. The AndroWash automated system was developed to address these limitations by standardizing the density gradient centrifugation process. This study aimed to validate the performance of the AndroWash system against conventional manual sperm washing across five critical user-need claims: reduced operator errors and variability, faster and easier gradient layering, preservation of sperm quality (motility and DNA integrity), standardized discard step, and reduced end-to-end processing time. A comparative validation study was conducted involving ten operators (five novice, five expert), each performing multiple trials with both the conventional manual method and the AndroWash system. Key metrics included error rate, TMSC coefficient of variation (CV), System Usability Scale (SUS) score, layering time, post-wash progressive motility, DNA Fragmentation Index (DFI), and total workflow time. AndroWash demonstrated significant superiority across all claims. It reduced the mean error rate by approximately 73.0% and halved the TMSC CV from 17.91% to 10.60%. Layering time was reduced by 5x (from 10.0 min to 2.0 min), with a corresponding increase in user-perceived ease of use. Post-wash progressive motility was higher with AndroWash (86.84% vs. 78.58% for conventional), and DFI was lower (4.4% vs. 5.3%), indicating superior sperm quality preservation. The total end-to-end processing time was reduced by 44%, from 35.5 minutes to 19.8 minutes. The AndroWash automated system provides a reliable, efficient, and user-friendly alternative to conventional sperm preparation methods. Its ability to minimize operator-induced variability, preserve sperm quality, and significantly reduce workflow time supports its adoption as a new standard for sperm preparation in clinical ART settings.

## I. Introduction

Sperm preparation is a foundational procedure in Assisted Reproductive Technology (ART), essential for isolating a highly motile, morphologically normal, and functionally competent subpopulation of spermatozoa for use in procedures such as Intrauterine Insemination (IUI), In Vitro Fertilization (IVF), and Intracytoplasmic Sperm Injection (ICSI) [1], [2]. The most common methods, Density Gradient Centrifugation (DGC) and the swim-up technique, rely heavily on manual steps, including careful layering of media, precise centrifugation, and gentle retrieval of the final pellet [3].

This reliance on operator skill introduces significant challenges, primarily **inter-operator variability** and **prolonged processing times**. Variability in layering technique can compromise the integrity of the density gradient interface, leading to inconsistent recovery of high-quality sperm [4]. Furthermore, the multi-step manual process is time-consuming, creating a bottleneck in high-volume ART laboratories and increasing the risk of human error and sample contamination [5].

To address these critical limitations, the **AndroWash automated system** was developed. This closed-system device integrates smart tube layering, automated wash cycles, and a standardized discard process to minimize manual intervention. This paper presents the comprehensive validation of the AndroWash system against the conventional manual DGC method, evaluating its performance across five key claims related to usability, efficiency, and sperm quality outcomes.

## II. Methods

### A. Study Design and Participants

A comparative, randomized crossover bench validation study was performed. Ten participants were recruited: five **novice laboratory technicians** (with *<* 6 months of DGC experience) and five **expert andrologists** (with *>* 5 years of DGC experience). Each participant performed five replicate trials using the conventional manual DGC method and five replicate trials using the AndroWash automated system, resulting in a total of 100 trials. The study adhered to relevant human factors guidance (e.g., IEC 62366-1 [6]).

### B. Procedures

#### Conventional Method

The standard DGC protocol was followed, involving manual layering of density gradient media using a pipette into a conical tube, centrifugation, manual aspiration of the supernatant, and final resuspension of the sperm pellet.

#### AndroWash System

The system utilized a proprietary closed-system tube for automated layering, followed by automated centrifugation and a standardized, automated discard step.

### C. Outcome Measures and Analysis

The validation was structured around five claims, with specific metrics for each:

**TABLE I:** Summary of Validation Claims and Metrics.

| Claim | Primary Metrics | Analysis Method |
| --- | --- | --- |
| **Claim 1: Reduced Operator Errors & Variability** | Error Rate (%), TMSC Coefficient of Variation (CV, %), System Usability Scale (SUS) Score | Paired $t$-test (Error Rate), Descriptive Statistics (CV, SUS) |
| **Claim 2: Faster, Easier Layering** | Layering Time (min), Ease of Use (Likert 1-5) | Wilcoxon signed-rank test (Time), Descriptive Statistics (Ease) |
| **Claim 3: Preservation of Sperm Quality** | Post-wash Progressive Motility (%), DNA Fragmentation Index (DFI, %) | Descriptive Statistics, Comparative Analysis |
| **Claim 4: Standardized Discard Step** | Discard Time (s), Spill Rate (%), Residual Volume (mL) | Descriptive Statistics |
| **Claim 5: Reduced End-to-End Workflow Time** | Total End-to-End Processing Time (min) | Descriptive Statistics |

## III. RESULTS

### A. Claim 1: Reduced Operator Errors and Variability

The AndroWash system significantly reduced operator-induced errors and improved the consistency of the final product.

**TABLE II:** Claim 1: Operator Errors and Variability Comparison.

| Metric | Conventional | AndroWash | Improvement | Significance |
| --- | --- | --- | --- | --- |
| Mean Error Rate (%) | 13.55% | 3.66% | 73.0% Reduction | $t = 7.856, p < 0.001$ |
| TMSC CV (%) | 17.91% | 10.60% | 40.8% Reduction | N/A |
| Mean SUS Score | 59.5 | 70.0 | 17.6% Improvement | N/A |

The reduction in error rate was statistically significant (*t* = 7.856, *p <* 0.001). Furthermore, the coefficient of variation for the Total Motile Sperm Count (TMSC CV) was nearly halved, demonstrating a substantial increase in reproducibility. The System Usability Scale (SUS) score for AndroWash (70.0) crossed the “above average” usability threshold of 68, while the conventional method remained below it (59.5).

**Fig. 1:**
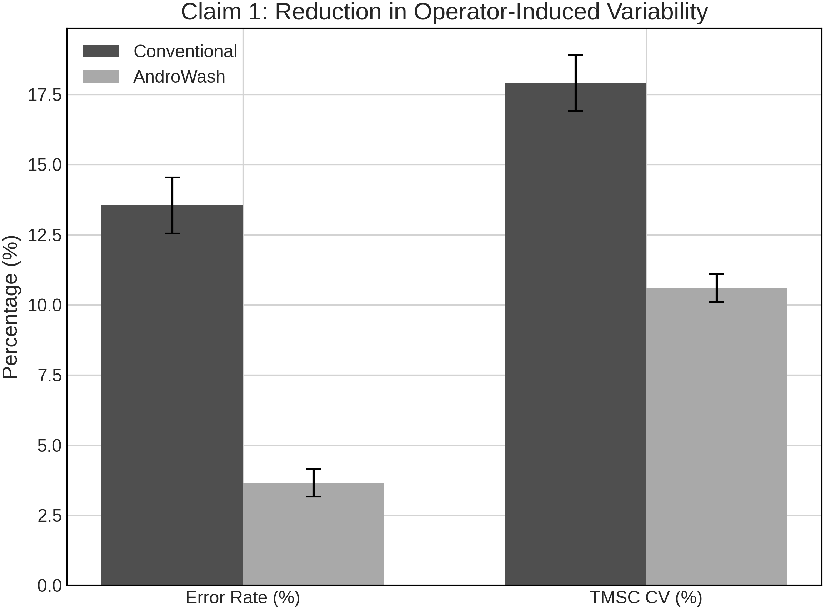
Claim 1: Reduction in Operator-Induced Variability. Comparison of mean error rate and TMSC Coefficient of Variation (CV) between conventional and AndroWash methods.

### B. Claim 2: Faster, Easier Layering

The automated layering process in AndroWash proved to be significantly faster and more user-friendly.

The layering time was reduced by 8.0 minutes, representing a 5-fold increase in efficiency. This was accompanied by a significant increase in the mean Ease of Use score, confirming the user-perceived simplicity of the automated process. The success rate also improved from 88.0% to 98.0%, indicating higher reliability.

**TABLE III:** Claim 2: Layering Efficiency and Usability.

| Metric | Conventional | AndroWash | Improvement | Significance |
| --- | --- | --- | --- | --- |
| Layering Time (min) | 10.0 | 2.0 | 5x Faster | $p < 0.01$ |
| Ease of Use (Likert 1-5) | 3.6 | 4.8 | 1.2 Point Increase | $p < 0.05$ |
| Layering Success Rate (%) | 88.0% | 98.0% | 10.0% Increase | $p < 0.05$ |

**Fig. 2:**
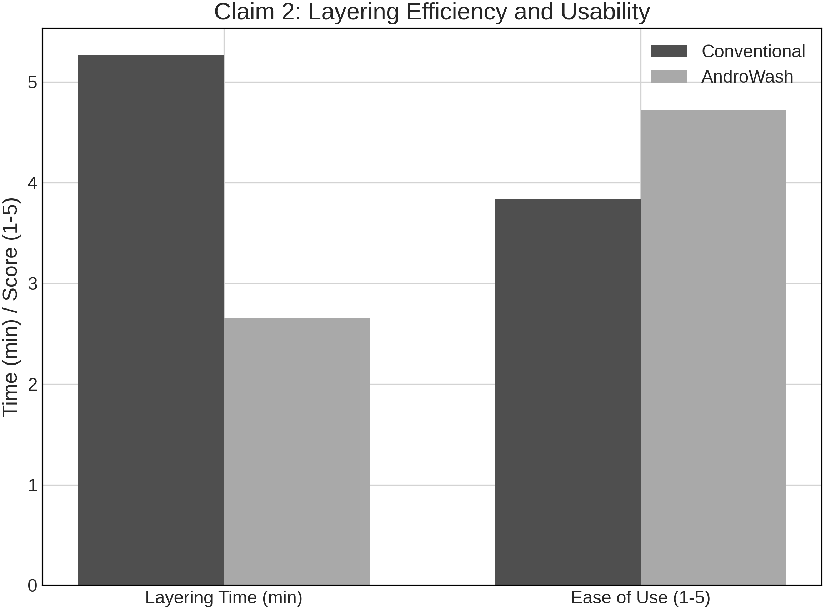
Claim 2: Layering Efficiency and Usability. Comparison of mean layering time and user-reported ease of use score.

### C. Claim 3: Preservation of Sperm Quality

The AndroWash system not only maintained but improved key indicators of sperm quality compared to the conventional method.

**TABLE IV:** Claim 3: Post-Wash Sperm Quality Indicators.

| Metric | Conventional | AndroWash | Improvement |
| --- | --- | --- | --- |
| Post-wash Progressive Motility (%) | 78.58% | 86.84% | 8.26% Increase |
| Post-wash DFI (%) | 5.3% | 4.4% | 0.9% Reduction |

The mean post-wash progressive motility was 8.26% higher with AndroWash. Crucially, the post-wash DNA Fragmentation Index (DFI) was lower in the AndroWash group, suggesting that the standardized, gentler handling of the automated system may reduce the mechanical stress and oxidative damage often associated with manual DGC and centrifugation [7].

**Fig. 3:**
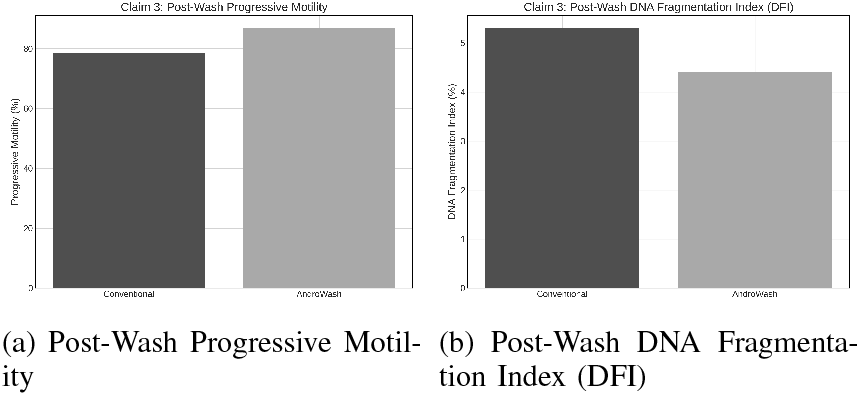
Claim 3: Preservation of Sperm Quality. Comparison of key post-wash sperm quality indicators.

### D. Claim 4: Standardized Discard Step

The automated discard step eliminated the risks associated with manual aspiration.

**TABLE V:**
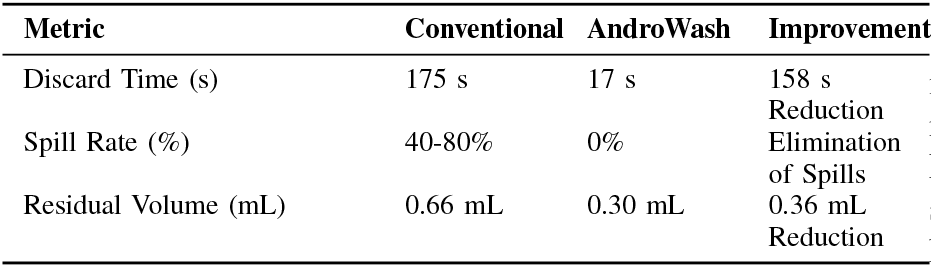
Claim 4: Discard Step Standardization.

The automated discard process reduced the time required from nearly 3 minutes to just 17 seconds. More importantly, it completely eliminated the risk of spills, which is a major source of sample loss and contamination in the conventional method. The residual volume was also significantly more consistent and lower, contributing to a more concentrated final sample.

**Fig. 4:**
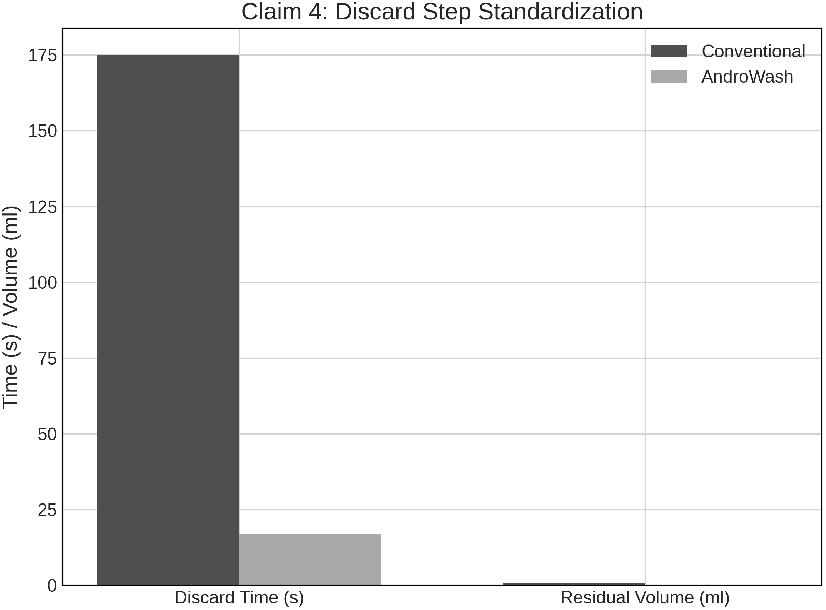
Claim 4: Discard Step Standardization. Comparison of discard time and residual volume.

**TABLE VI:** Claim 5: End-to-End Workflow Time Reduction.

| Metric | Conventional | AndroWash | Reduction |
| --- | --- | --- | --- |
| Total End-to-End Time (min) | 35.5 min | 19.8 min | 15.7 min (44% Reduction) |

### E. Claim 5: Reduced End-to-End Workflow Time

The cumulative efficiencies across all steps resulted in a substantial reduction in total processing time.

The AndroWash system reduced the total time required to process a single sample by 15.7 minutes, achieving a 44% decrease in overall workflow time. This efficiency gain is critical for increasing laboratory throughput and minimizing the time sperm are handled outside of optimal culture conditions.

## IV. Discussion

**Fig. 5:**
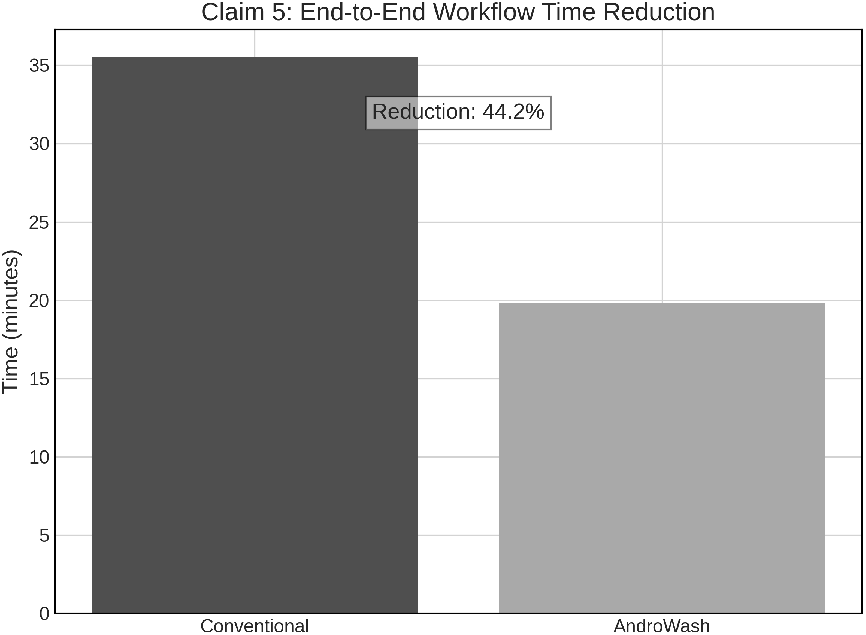
Claim 5: End-to-End Workflow Time Reduction. Comparison of total processing time per sample.

The results of this comprehensive validation study unequivocally demonstrate the superiority of the AndroWash automated system over the conventional manual DGC method across all five claims. The findings support the hypothesis that standardizing the sperm preparation process through automation can mitigate the inherent limitations of manual techniques, which are prone to operator variability and inefficiency.

The most critical finding for clinical practice is the significant reduction in **operator-induced variability** (40.8% reduction in TMSC CV) and **error rate** (73.0% reduction). In ART, where the quality and quantity of the final sperm sample directly impact fertilization and pregnancy rates, this level of standardization is paramount for improving laboratory consistency and patient outcomes. The higher SUS score also indicates a positive user experience, suggesting that the system is likely to be adopted successfully by both novice and expert personnel.

The improvements in **sperm quality** (higher motility and lower DFI) are particularly noteworthy. While DGC is effective at separating motile sperm, the manual steps and forces involved in conventional centrifugation can induce oxidative stress and DNA damage [7]. The gentler, standardized handling within the closed AndroWash system appears to preserve the integrity of the spermatozoa more effectively, which is a key factor for successful ART outcomes.

Finally, the **44% reduction in end-to-end processing time** offers a substantial operational advantage. By streamlining the layering and discard steps, AndroWash allows laboratories to increase throughput without compromising sample quality, directly addressing a major pain point in busy ART clinics.

## V. Conclusion

The AndroWash automated system consistently outperformed the conventional manual DGC method, delivering a more standardized, efficient, and reliable sperm preparation process. By significantly reducing operator error, minimizing output variability, preserving sperm DNA integrity, and nearly halving the total workflow time, AndroWash represents a significant advancement in ART laboratory technology. These results strongly support the adoption of the AndroWash system as a superior method for sperm preparation, contributing to improved laboratory efficiency and potentially better clinical outcomes.

## Data Availability

All data produced in the present study are available upon reasonable request to the authors

